# Experiences and needs of homebound patients and caregivers of oral nutritional management after pancreaticoduodenectomy: a qualitative study using Knowledge-Attitude-Practice theory

**DOI:** 10.1101/2025.06.14.25329463

**Authors:** Cui Gao, Yongping Gao, Qiuge Qiao, Yehong Kong, Ci Dong

**Affiliations:** Department of General Surgery I, The Second Hospital of Hebei Medical University; Department of Nursing, The Second Hospital of Hebei Medical University, Luquan Campus; Department of General Surgery I, The Second Hospital of Hebei Medical University, Luquan Campus; Department of Neurology, The First Hospital of Hebei Medical University

**Keywords:** Pancreaticoduodenectomy, Knowledge-Attitude-Practice Theory, Transoral Nutritional Management, Qualitative study

## Abstract

**Objective:** To understand the real-life experiences and needs of homebound patients during oral nutritional management after pancreaticoduodenectomy to inform the construction of targeted intervention programs.

**Methods:** Semi-structured interviews were conducted with a total of 25 individuals who were homebound patients and primary caregivers after pancreaticoduodenectomy, and themes were analyzed and summarized using directed content analysis.

**Results:** Three themes and seven sub-themes were identified. These included Knowledge-lack of knowledge about transoral nutritional management, existence of transoral nutritional misconceptions, and active search for knowledge related to transoral nutrition;Attitude-multiple physical-psychological distress leading to low confidence (distress of GI discomfort symptoms, distress of negative psychological emotions after eating), perceived benefits and gains; Practice-lack of action threads, active exploration of appropriate dietary behaviors.

**Conclusions:** The experiences and needs of patients and caregivers at home after pancreaticoduodenectomy in the process of transoral nutritional management are diverse. Nurses should do a good job of post-discharge follow-up, pay attention to the needs and psychological status of patients and caregivers in the process of transoral nutritional management, and focus on identifying barriers to transoral nutritional management in order to provide professional and personalized guidance and support.

## 1 Introduction

Pancreaticoduodenectomy (PD) is a standard procedure for the treatment of various diseases around the head and jugular abdomen of the pancreas and is one of the common procedures in general surgery. The surgical procedure involves resection of multiple organs (part of the stomach, duodenum, neck of the pancreatic head, common bile duct, gallbladder, and part of the jejunum) and reconstruction of the digestive tract (pancreatico-enteric anastomosis, biliary-enteric anastomosis, and gastrointestinal anastomosis)^1^. Due to the immunometabolic characteristics of the primary disease , preoperative nutritional risk and malnutrition, the impact of surgery on the structural and functional changes of the digestive system, surgical complications, and postoperative tumor-related adjuvant therapies, approximately 52% to 88% of patients undergoing surgery are at moderate to severe risk of malnutrition^2^. And nutritional status is strongly associated with disease treatment efficacy, risk of recurrence and clinical outcomes^3^.

As pancreaticoduodenectomy continues to develop and become more sophisticated in terms of nutritional supportive therapy, surgical techniques, and rapid postoperative rehabilitation^4–6^, which greatly reduces the length of hospitalization, patients are in greater need of long-term post-discharge nutritional guidance and intervention. Some studies have shown that the proportion of patients readmitted to the hospital due to malnutrition after pancreaticoduodenectomy reaches 16% ^7,8^, and 90% of cases occur in the community setting^9^, and recovery of postoperative nutritional status takes at least 3 months or even longer, even with close monitoring and support ^10^.

The five-step treatment model of malnutrition proposed by the Professional Committee of Tumor Nutrition and Supportive Therapy of the Chinese Anti-Cancer Association^11^ points out that transoral nutrition is the preferred route of nutritional therapy. When post-pancreaticoduodenectomy patients are discharged from the hospital and enter the stage of home rehabilitation, the intake of nutrients by the oral route is basically restored, and the task of home rehabilitation is mainly undertaken by the patients themselves and their caregivers. Several studies have shown^12,13^ that the caregiving experience and needs of family caregivers have a direct impact on the quality of life of cancer patients, so the experience and needs of caregivers should not be ignored. Studies have confirmed^14,15^ that oral nutritional interventions can effectively improve dietary compliance, prevent malnutrition and increase food intake in pancreaticoduodenectomy patients. Effective health education can not only improve the patients’ understanding of the disease, but also make the patients consciously cooperate with the treatment^16^. Knowledge-Attitude-Practice (KAP) is a theoretical model for changing human health-related behaviors, which divides the change of human behaviors into three consecutive processes of acquiring knowledge, generating beliefs and forming behaviors, i.e., knowledge-attitude-practice^17^. This study intends to analyze the real experiences and needs of homebound patients and caregivers in the process of oral nutrition management after pancreaticoduodenectomy from the aspects of knowledge, beliefs, and behaviors through descriptive qualitative research methodology, using the KAP model as a theoretical framework, with the aim of improving the nutritional status and quality of life of the patients, and at the same time to provide reference for the development of intervention programs for oral nutrition management in homebound patients after pancreaticoduodenectomy.

## 2 Methods

### 2.1 Design

A descriptive qualitative research methodology was used to conduct face-to-face, semi-structured, in-depth interviews with eligible post-pancreaticoduodenectomy homebound patients and caregivers to gather information about their experiences and needs during transoral nutritional management.

### 2.2 Participants and setting

Purposive sampling method was used to select homebound patients and primary caregivers after pancreaticoduodenectomy in the Department of General Surgery I of a tertiary hospital in Hebei Province from February to July 2024 as the study subjects based on the information of the patients’ gender, marital status, literacy level, type of pathology, and postoperative time by using the maximum difference sampling strategy. Inclusion criteria: ① post-pancreaticoduodenectomy patients, age ≥ 18 years old; ② good language and communication skills; ③ informed consent, voluntary participation in this study. Exclusion criteria: (1) tube-feeding patients; (2) mental illness or other serious illnesses that prevented the completion of the study. Inclusion criteria for caregivers: (1) family members of post-pancreaticoduodenectomy patients who are the primary caregivers at home and are ≥ 18 years old; (2) good language and communication skills; (3) informed consent and voluntary participation in the study. Exclusion criteria: primary caregivers who need to pay for care. The sample size was based on the principle of saturation of the interview data and the absence of new themes.

### 2.3 Interview Guide

A semi-structured interview guide was developed based on the literature review and KAP theory to ensure that the questions were effective in eliciting responses consistent with the study objectives. The first draft of the interview guide was validated by two experts, including a nutritional nurse specialist and a pancreatic surgery chief nurse practitioner, to ensure that the questions were appropriate. To further ensure the comprehensiveness of the interview guide, pre-interviews were conducted, including two homebound patients after pancreaticoduodenectomy and two caregivers, and the interview guide was revised. The final interview guide is shown in Table 1.

**Table 1.** Interview guide.

| Interviewees | Interview Guide |
| --- | --- |
| **Patient** | ①How did you manage your transoral diet and nutrition during your post-surgical home stay?<br>②What changes did you experience in your trans-oral diet and nutrition during your post-surgical home stay? How did you cope?<br>③What problems and difficulties did you experience with your transoral diet and nutrition during your post-surgical home stay? How did you manage these problems and difficulties?<br>④What help and support would you like to receive to assist you in managing your Transthoracic Diet and nutrition? Can you be more specific? |
| **Caregiver** | ①How do you manage your patients in terms of transoral diet and nutrition during the post-surgical home stay?<br>②What are some of the changes that occur during the post-surgical homebound period for patients in terms of trans-oral diet and nutrition? How do you help patients cope?<br>③What problems and difficulties have you encountered in caring for your patient's trans-oral diet and nutrition while at home after surgery? How did you resolve these problems and difficulties?<br>④What help and support would you like to receive to assist yourself in the management of your patients' transoral diet and nutrition? Can you be more specific? |

### 2.4 Data collection

Data were collected using a semi-structured in-depth interview method . The interviewer was a nurse who had worked in pancreatic surgery for 15 years. Patients and caregivers were screened based on inclusion and exclusion criteria prior to the start of the interview. The interviewer established contact with the respondent in advance to briefly introduce and build trust, and set a time and place for the interview. Respondents were informed of the purpose, methodology, and use of the study, fully informed and signed a consent form prior to the interview, and a general information questionnaire was distributed to collect information about their age, gender, relationship to the patient, literacy level, place of residence, and occupation.The interviewers had completed learning and training modules on qualitative research methods before the interviews to ensure that the interview process was reasonable and standardized. The interviews were conducted in a quiet and private consultation room. The interviews were recorded with the interviewees’ consent. By using various techniques such as careful listening and feedback and raising appropriate follow-up questions, the interviewers proceeded with the in-depth interviews and encouraged the interviewees to express their thoughts and feelings freely. The interviewees’ facial expressions and body movements were recorded in a timely manner without using guiding or suggestive language. Each interview lasted for 15–40 min.

### 2.5 Rigor

Purposive sampling of participants with different demographic characteristics to ensure transferability. The interviewers remained neutral throughout the interviews and did not express their personal opinions or judgments. When unclear statements or feelings arose, the interviewers sought clarification from the participants during the interview. In the data analysis process, the original statements of the interviewees were acknowledged without personal interpretations or opinion. Data analysis was conducted to reach consensus with co-researchers on the categories and subcategories, enhancing the credibility and confirmability of the findings. After the data analysis, the textual data were given to the interviewees for validation to ensure the stability of the results.

### 2.6 Data analysis

The audio recordings were transcribed textually, labeled with nonverbal information, and data analysis was conducted by two researchers in pairs within 24h after the interviews were completed. In this study, the data were analyzed using the directed content analysis method^18^, which consists of 3 main phases: the initial data preparation phase, the organizational coding phase, and the reporting of results. The specific steps are as follows: the researcher repeatedly and carefully read the original interview data until she had an in-depth understanding of its overall content; according to the purpose of this study, KAP theory was used as a framework to determine the categories of the units of analysis; open coding was used to label important ideas and concepts in the data, and similar codes were grouped into the corresponding categories to form themes and sub-themes. Coding was done using a combination of manual coding and Nvivo 11.0 software.

### 2.7 Ethical considerations

The study was reviewed by the Hospital Ethics Committee (IRB No. 2024-R023), and the subjects provided informed consent and voluntarily participated in the study.

## 3 Results

### 3.1 Participants’ characteristics

A total of 12 patients (P1-P12) and 13 caregivers (C1-C13) who were homebound after pancreaticoduodenectomy were interviewed for this study, and general information about the patients and caregivers is shown in Tables 2 and 3.

**Tables 2.** General patient information (n=12)

| No. | Gender | Educational | Marital status | Residence | Pathologic type | Postoperative time (months) |
| --- | --- | --- | --- | --- | --- | --- |
| P1 | Female | Primary school | Married | Rural | Bile ducts middle and low differentiated adenocarcinoma | 11 |
| P2 | Male | High school | Widowed | Rural | Duodenal adenomyoma | 7 |
| P3 | Female | High school | Married | Urban | Gastrointestinal mesenchymal tumor | 5 |
| P4 | Female | Bachelor's degree | Unmarried | Urban | Solid pseudopapillary tumor of the pancreas | 6 |
| P5 | Male | Middle school | Married | Rural | Gastrointestinal mesenchymal tumor | 5 |
| P6 | Female | Primary school | Married | Rural | Medium to low differentiated carcinoma of the pancreas | 10 |
| P7 | Male | High school | Married | Urban | Ductal adenocarcinoma of the pancreas low to moderately differentiated | 5 |
| P8 | Female | Primary school | Married | Rural | Highly differentiated adenocarcinoma of jugular abdomen | 1 |
| P9 | Male | Primary school | Married | Rural | Low to moderately differentiated adenocarcinoma of the jugular abdomen | 4 |
| P10 | Female | Primary school | Married | Urban | Plasma cystadenoma of pancreas | 3 |
| P11 | Female | Middle school | Married | Rural | Hypo-differentiated adenocarcinoma of duodenal papilla | 7 |
| P12 | Male | Middle school | Married | Urban | Middle and low differentiated carcinoma of the pancreas | 5 |

**Tables 3.** General information about caregivers (n=13)

| No. | Gender | Educational | Residence | Relationship with patients |
| --- | --- | --- | --- | --- |
| C1 | Female | Bachelor's degree | Urban | child |
| C2 | Male | Bachelor's degree | Urban | child |
| C3 | Male | Middle school | Rural | child |
| C4 | Male | Junior college | Urban | child |
| C5 | Male | Middle school | Rural | child |
| C6 | Female | Middle school | Rural | partner |
| C7 | Female | Middle school | Rural | partner |
| C8 | Male | Middle school | Urban | partner |
| C9 | Male | Primary school | Rural | partner |
| C10 | Female | Junior college | Rural | child |
| C11 | Female | Middle school | Rural | partner |
| C12 | Female | Bachelor's degree | Urban | child |
| C13 | Male | Bachelor's degree | Urban | child |

### 3.2 Results of thematic refinement

#### Theme 1: Knowledge

##### 1. Lack of knowledge of transoral nutritional management

After discharge from the hospital, patients are physically weak, and most of their caregivers are responsible for their diet and part of their daily life. The caregiver’s knowledge of transoral nutritional management and the patient’s own nutritional knowledge directly affect the patient’s postoperative recovery and nutritional intake, and most of the interviewees have vague concepts of nutrition. C7: “We are a farmer in the countryside and we don’t know anything about nutrition, but we should eat eggs and meat. Eating eggs and meat is the only way to be nutritious!” P7: “We just drink porridge and thin food every day, but it’s not nutritious! If I want to eat something nutritious, I don’t know what to eat.” C9: “We don’t have to be so particular at home, we just give her stewed chicken and beef, and she eats it every now and then.”

#### 2. Misconceptions about transoral nutrition

Many post-pancreaticoduodenectomy patients and caregivers have a relatively poor nutritional knowledge base due to age, differences in educational backgrounds, limited economic resources, and restricted access to information, and are poorly utilized in practice. This is reflected in their generally low level of transdermal nutritional knowledge and is often accompanied by erroneous dietary concepts and behavioral habits. P5: “I don’t drink milk, I’ve heard that drinking milk is bloating and diarrhea, and it’s not well digested.” C9: “I heard people say leeks, celery do not allow to eat, that is not good for digestion, fish, shrimp did not dare to let her eat it, these are hairy, eat the wound growth is not good.” C6: “Basically are fluids, I stewed ribs, do not dare to let him eat meat, just take the ribs soup for him to cook fine noodles to eat, cooked rotten paste, good digestion and nutrition.”

#### 3. Actively seeking knowledge about transoral nutrition

Most post-pancreaticoduodenectomy patients and caregivers also actively seek information and guidance on transoral nutritional management using a variety of channels, such as television, books, surfing the Internet on Baidu, short video accounts, public numbers, online forums, and so on. However, the information obtained is mostly fragmented content, and the authority and validity are difficult to ensure. P2: “I saw on TV that it is better to let eat more vegetables, and it is better to eat the green ones, and try not to eat fried things.” P3: “I read online that it is better to eat more chicken, which is high in protein.” C8: “Sometimes I look it up online, but it basically says the same thing, and it’s not too obvious to try and try.” C12: “I saw on Jitterbug that eating sea cucumber is good, and there are videos teaching how to make sea cucumber, sea cucumber porridge, fried sea cucumber with scallions, and I made it for him in different ways, and he quite loves it.”

### Theme 2: Attitude

#### 1. Multiple physical and psychological distress leads to low confidence

##### (1) Trouble with digestive discomfort symptoms

Pancreaticoduodenectomy involves the resection of multiple organs and the reconstruction of the digestive tract, which has a great impact on the patient’s diet and digestive function after surgery. In addition to leading to impaired gastrointestinal function of patients, it also affects the internal and external secretion function of the pancreas, leading to lipid absorption disorders and glucose metabolism disorders in patients after surgery^19^. Almost all interviewees expressed the existence of digestive discomfort symptoms during the interview and lost confidence in disease recovery.P5: “Sometimes I don’t dare to eat at night, my stomach swells when I eat, and I have acid reflux and hiccups, am I not recovering well?” P2: “My stomach gurgles, I have frequent bowel movements and urinate a lot.” C10: “I always feel that gas is flowing in my belly, and it hurts here at the top of my heart.” C7: “After eating something, he feels bloated in his stomach and can’t rest even if he wants to.” C8: “If he eats a little bit of greasy food, he feels uncomfortable and nauseous.” After pancreaticoduodenectomy, the physiological structure and function of the gastrointestinal tract of the patient have changed, which is mainly manifested in the obvious symptoms of the digestive tract after eating.

##### (2) Negative Psychological Emotional Distress After Eating

The patient’s anxiety over the fear of eating triggering various uncomfortable GI symptoms manifests itself in a reluctance to eat or a fear of eating.P3: “I don’t dare to drink milk, I’m afraid it’s indigestible.” P7: “I want to eat, but it’s hard when I eat.” C5: “At first she didn’t dare to eat anything and couldn’t eat, she was afraid of discomfort, then she didn’t dare to eat.” C7: “He said he ate too much but he didn’t realize it was that uncomfortable, so he didn’t dare to eat more anymore.” At the same time, a series of chain reactions triggered by reduced eating, such as general weight loss or no gain, weakness, powerlessness, lack of spirit, etc., also made many patients and caregivers worried.C2: “She doesn’t eat much, she always feels weak, I also want her to eat more, but she can’t eat it! There’s nothing I can do about it.” P3: “I just don’t gain weight. I would feel better if I gained a few pounds, but I don’t feel like I’m recovering well.” Some patients reported that they ate because of external pressure, either due to the need to recover from the disease or pressure from their caregivers.P6: “I just have to eat when it’s time to eat, I’m not really hungry.” P7: “She’s there telling me ‘have a dish of this, have a dish of that’ and I don’t want to eat (waving her hand).” C7: “Every time he was forced to eat like that, he ate.” There were also patients who expressed the need and desire for food, but the conflict between eating and discomfort after eating filled them with conflict and helplessness.P9: “I wanted to eat meat the other day, and I ate some ribs, and some beef, and it made me hard to eat.” P3: “I felt able to eat, but then I started to feel bad after eating, and now it makes me afraid to eat.”

#### 2. Perceived benefits and gains

Most of the post-pancreaticoduodenectomy patients found their knowledge and understanding of transoral nutrition significantly improved during their subsequent treatment and recovery, and they began to pay attention to the nutrient content of food, learned to differentiate between healthy and unhealthy food, and experienced first-hand the benefits of transoral nutritional management on their physical recovery.P11: “I used to eat less vegetables, and when I was discharged from the hospital The nurse told me to eat more fresh vegetables, I now eat broccoli, carrots, tomatoes, whatever, fresh vegetables in season anyway, and my stools are normal and I am not constipated anymore.” P9: “Now you can’t eat blindly, you can’t just eat whatever you want, it has to be well digested and nutritious.” C1: “Originally she was a non-spicy person, she couldn’t eat without spicy food, now she doesn’t dare to eat spicy food, saying that spicy food is not good for digestion.” Meanwhile, the support and encouragement of caregivers can not only enhance patients’ confidence in overcoming the disease, but also better help patients with transoral nutritional management.P12: “When I first went back to the hospital, in order to make me eat nutritious food that is good for digestion, my children used the food processor to make all kinds of rice pudding for me, which is quite tasty.” P10: “The last review said that my protein was low, so my partner said that we had to eat well and let me drink protein powder, and last week’s review said that my protein was normal, so my partner can take credit for that.”

### Theme 3: Practice

#### 1. Lack of action threads

Some post-pancreaticoduodenectomy patients or caregivers reported that they ate a single type of food every day and had certain dietary preferences, and that their daily dietary habits were not easy to change and lacked clues for action, so they remained fixated on traditional nutritional perceptions.C2: “My parent doesn’t eat meat, she just eats a lot of veggies. just eat more vegetarian food.” C3: “Every time she eats, she eats more staple food, steamed buns, some vegetables, rice porridge, and little milk, and she doesn’t drink it even when it’s spoiled.” P8: “Every time I eat I like to match a little pickled radish or small pickles, eat the mouth with flavor, they all say you can not eat these pickled foods, not good for the body, but I formed the habit, do not eat some pickles, always feel the rice does not smell good.”

#### 2. Active exploration of appropriate dietary behaviors

Patients or caregivers actively engage in oral nutrition management by adjusting intake, trying different types of food, or changing the way food is processed to reduce GI symptoms and promote recovery, and gradually establish appropriate eating behaviors through repeated experimentation in practice.C9: “When I first came home I gave her a set time to eat, which was about 3 clock a meal. If she wasn’t hungry she had two sips less, if she was hungry she had two sips more, that’s how I scheduled it for her.” P1: “If her stomach gets sick, she eats less for a couple of days, and then when she gets better, she eats more, and I try to make sure that I don’t eat too much.” C8: “I bought that kind of deep-sea fish from the supermarket and steamed it for her to eat. According to my understanding, it is quite soft when steamed, so it should be easy to be absorbed after eating it.” P9: “Now I probably have some idea of what to eat, unlike in the past when I didn’t know what to eat, but I just try. Especially this amount of food for meals, it’s all felt out.” Some patients manage their dietary behavior by learning from the successes of other patients.C4: “The last time we came for chemotherapy, the patient in the next bed had the same disease as my parent, and she usually ate well and had good nutrition, saying that she needed to eat more often and exercise more often in order to build up her resistance. After she came home my parent was quite active and said she wanted to learn from others.”

## 4 Discussion

### 4.1 Enhancing knowledge of transoral nutritional management to improve nutritional literacy among homebound patients and caregivers after pancreaticoduodenectomy

This study found that the lack of knowledge about transoral nutritional management and the existence of transoral nutritional misconceptions among post-pancreaticoduodenectomy patients and caregivers will directly affect patients’ eating behavior. A survey on the status of nutritional knowledge, beliefs and behaviors of digestive system tumor patients showed^20^: 76.8% of patients’ nutritional knowledge was at a poor level, and at the same time, there were still traditional misconceptions about some nutritional knowledge; 64.2% of patients’ nutritional attitudes were at a positive level, but only a few of them were willing to take the initiative to seek nutritional knowledge or consult with others. Therefore, healthcare professionals can make full use of this feature by using digital health-based platforms such as “Internet +”, the Internet of Things, and smartphone apps to regularly push relevant oral nutrition knowledge to homebound patients and their caregivers after pancreaticoduodenectomy and to set up online counseling services. Through these initiatives, patients and their caregivers are guided to actively learn more about nutrition, establish correct dietary concepts, and develop healthy eating behaviors^21^. In addition, the level of nutritional literacy of patients or caregivers is closely related to sociodemographic factors such as literacy, gender, age, and economic status. In general, more educated or younger patient groups are more active in learning about the disease and have more diverse access to medical information. Of course, the lower nutritional literacy level of oncology patients is also related to the limited nutritional knowledge of healthcare personnel, the single way of acquiring knowledge during hospitalization, and the lack of continuity of care^22^. Healthcare professionals are the key channel for patients to obtain oral nutritional education and dietary guidance, so healthcare professionals with good nutritional literacy play an important role in the nutritional awareness of homebound patients and caregivers after pancreaticoduodenectomy. This emphasizes the need for healthcare professionals to first strengthen their nutritional expertise to ensure that they are able to provide accurate and scientific guidance to patients. Second, given the diverse social background of patients and caregivers, healthcare professionals should flexibly use online and offline methods to customize personalized nutrition education and dietary guidance programs to meet different needs. Furthermore, by increasing the intensity of education and focusing on the practicality of the content and the diversity of the forms, we aim to profoundly influence the nutritional concepts of patients and caregivers, so that they fully recognize the importance of oral nutrition and its positive impact on recovery, and at the same time be alert to and correct the nutritional cognitive misunderstandings. Ultimately, it guides patients and caregivers to form healthy and scientific eating habits, and realizes early education, timely correction and effective prevention of nutritional problems.

### 4.2 Timely interventional interventions to effectively address post-feeding discomfort in homebound patients after pancreaticoduodenectomy

Some studies have shown that the symptom burden of patients after pancreaticoduodenectomy is high and is most prominent in the 6 weeks after surgery^8^. Most of the patients in this interview reported eating only 1/3 to 1/2 of their preoperative intake, accompanied by nausea, vomiting, loss of appetite, abdominal pain and bloating, dyspepsia, and other discomfort symptoms. Therefore, medical staff should intervene early to intervene, detect the problem in time and implement relevant therapeutic measures to minimize its impact on patients’ diet and impede the normal intake of nutrition. At present, clinical medical workers usually use the inquiry method to assess the appetite of tumor patients, which is subjective and difficult to quantify. As mentioned in the Chinese Guidelines for the Integrated Diagnosis and Treatment of Cancer (CACA), tools such as the Anorexia/Hypoglossia Evaluation Scale^23^, the Appetite Symptom Questionnaire for Tumor Patients, and the Appetite Scale^24^ can be used to quantitatively evaluate the appetite of tumor patients. For post-pancreaticoduodenectomy patients with loss of appetite, glucocorticoids, progestins and other pro-appetizing drugs are commonly used, which can be supplemented with a combination of alternative Chinese medicine therapies such as moxibustion^25^, acupoint compresses^26^, soups^27^, or non-pharmacological interventions such as psychotherapy. The diet of patients with nausea and vomiting can be small, frequent and well-digested. Some gastric power promoting drugs such as metoclopramide and domperidone can improve gastric fullness and satiety in tumor patients. Oral pancreatic enzyme preparations can treat steatorrhea and abdominal cramps caused by pancreatic exocrine insufficiency^28^.For chemotherapy-induced nausea and vomiting, antiemetic drugs can be used as prescribed by the doctor; for milder cases, patients can be relieved by music therapy^29^, muscle relaxation training^30^, and aromatherapy^31^. If altered taste sensation is present, patients can try rinsing their mouth with warm boiled water or lightly salted water before meals, and it is recommended to reduce the intake of foods that may exacerbate the sensation of bitterness and accordingly increase the intake of acidic foods and foods with strong flavors in order to enhance the eating experience. In addition, caregivers can be advised to change food types frequently, carefully mix and match the colors of dishes, and adopt diversified cooking methods to enhance visual, olfactory, and gustatory stimulation to stimulate the appetite of post-pancreaticoduodenectomy patients, and improve their acceptance of and compliance with their diets.

### 4.3 Multifaceted guidance to build positive attitudes and confidence in transoral nutritional management in homebound patients after pancreaticoduodenectomy

This study found that post-pancreaticoduodenectomy patients faced gastrointestinal discomfort due to eating during the home rehabilitation phase, which triggered different degrees of negative psychological emotions, a phenomenon that significantly weakened patients’ self-confidence and motivation to implement transoral nutritional management strategies. Studies^32^ have shown that there is a bidirectional regulation between diet and mental health, and that negative emotions affect patients’ eating behaviors and nutrient absorption, thus adversely affecting patients’ quality of life and disease prognosis. Therefore, constructing and maintaining a positive mindset is an indispensable prerequisite for the successful implementation of oral nutritional management strategies in homebound patients after pancreaticoduodenectomy. At the same time, strengthening an individual’s self-efficacy and belief in his or her ability to successfully implement and adhere to this nutritional management program is a core intrinsic driver for sustaining transoral nutritional management. Studies have shown that patients with high self-efficacy are able to cope with the disease with optimism, have greater stress tolerance and psychological stability, and are able to withstand the negative stress associated with the disease^33^. It is suggested that healthcare professionals should focus on the self-efficacy of homebound patients after pancreaticoduodenectomy and the presence of negative psychological emotions when performing transoral nutritional management, and encourage patients to share their inner feelings by organizing expressive writing and supportive communication groups to guide them to manage and regulate their own emotions effectively^34,35^. In addition, healthcare professionals should be committed to building bridges of positive mindset in the rehabilitation process of homebound patients after pancreaticoduodenectomy, and assist patients in dispelling negative psychological emotions and regaining confidence through the implementation of psychological interventions such as positive stress reduction therapy and motivational interviewing. This process aims to stimulate patients’ intrinsic potential, learn from the successes of others, and deeply realize the importance of adhering to transoral nutritional management to improve nutritional status and promote the recovery process, thus strengthening their sense of self-efficacy. At the same time, by clearly explaining the possible negative consequences of neglecting transoral nutritional management strategies, patients are guided to reexamine and adjust their attitudes toward the disease, so that they can accurately grasp the balance between the benefits and potential obstacles, and realize the optimization of self-management^36^. In addition, the self-drive and self-monitoring abilities of homebound patients after pancreaticoduodenectomy can be improved by setting appropriate goals, monitoring weight changes, and recording dietary diaries to promote the adoption of healthy and nutritious eating behaviors.

### 4.4 Fully mobilizing multidimensional social support systems and reinforcing the positive impact of important players on the oral nutritional behavior of homebound patients after pancreaticoduodenectomy

Social support can come from a variety of sources such as family, patients, and social organizations. For homebound patients after pancreaticoduodenectomy, good social support plays a crucial role in adapting to adversity, adjusting their mindset, and positively coping with the challenges of the disease, and can also provide strong support for their adherence to transoral nutritional management behaviors. From the dimension of family support, patients have the longest contact time with their caregivers. Therefore, when family support is increased, it can maximize the ability to help homebound patients after pancreaticoduodenectomy to adjust their mindset and enhance their confidence in actively fighting the disease. In view of this, healthcare professionals must pay attention to supportive education for caregivers, which can be used in ways such as binary coping interventions^37^ to actively mobilize caregivers to participate in the patient’s transoral nutritional management, thereby increasing the level of family support. At the same time, healthcare professionals should also encourage patients to express their needs and emotions in order to gain more understanding and support from their families. This requires healthcare professionals to not only strengthen transoral nutrition education and dietary guidance for patients, but also pay attention to the corresponding education for their caregivers, increase the relevant knowledge reserves, improve the understanding of the relevant risks, and effectively respond to a series of symptoms that may occur in the process of transoral nutrition for home-bound patients after pancreaticoduodenectomy, which may affect the eating process. Caregivers should not only guide and encourage patients in thought and give them emotional support, but also strictly monitor and supervise patients’ transoral dietary behaviors in action, so as to play the role of positive incentives. At the level of patient support, patients should be motivated to join patient associations and disease communication groups to share their experiences of transoral nutritional management with other patients or to talk about psychological disturbances. In this way, patients are assisted to build a solid patient support network, which in turn relieves their psychological pressure and eliminates their sense of isolation.In addition, healthcare professionals are also an important social support force. Departmental administrators can organize a multidisciplinary team to carry out comprehensive transoral nutritional management and follow-up for homebound patients after pancreaticoduodenectomy. Providing patients with practical and effective continuity of care services, continuous psychological support and counseling and guidance on transoral nutrition will help patients cope with the multiple physical and psychological challenges and improve their ability to manage transoral nutrition.

## Conclusions

Using KAP as a theoretical framework, this study conducted semi-structured interviews on the experiences and needs of homebound post-pancreaticoduodenectomy patients and caregivers in the process of transoral nutritional management, and three themes and seven sub-themes were distilled, including Knowledge-lack of knowledge about transoral nutritional management, existence of transoral nutritional misconceptions, and active search for knowledge related to transoral nutrition; Attitude -multiple physical-psychological distress leading to low confidence (distress of GI discomfort symptoms, distress of negative psychological emotions after eating), perceived benefits and gains; and practice-lack of action threads, active exploration of appropriate dietary behaviors. The experiences and needs of homebound patients and caregivers after pancreaticoduodenectomy in the process of transoral nutritional management show diversity, and nurses should do a good job of post-discharge follow-up to pay attention to the needs and psychological status of patients and caregivers in the process of transoral nutritional management, with a focus on identifying obstacles that affect transoral nutritional management in order to provide professional and personalized guidance and support.

## Limitations

The subjects included in this study were from only one tertiary care hospital, the results may have some limitations, and future multi-center clinical studies are still needed to validate the results of this study.

## Data Availability

The data that support the findings of this study are available on request from the corresponding author. Data are not made public due to privacy and ethical restrictions.

## CRediT authorship contribution statement

**Cui Gao:** Conceptualization, Methodology, data collection, transcription and summarization, Writing-Original draft, Writing-Review and editing.

**Yongping Gao:** Conceptualization, data collection, transcription and summarization.

**Qiuge Qiao:** Conceptualization, Methodology, data collection, transcription and summarization.

**Yehong Kong:** Conceptualization, Methodology, data collection.

**Ci Dong:** Conceptualization, Methodology, Writing-Review and editing. All authors had full access to all the data in the study, and the corresponding author had final responsibility for the decision to submit for publication. The corresponding author attests that all listed authors meet authorship criteria and that no others meeting the criteria have been omitted.

## Ethics statement

This study was approved by the Research Ethics Committee of the Second Hospital of Hebei Medical University (IRB No. 2024-R023),All participants provided written informed consent.

## Declaration of Generative AI and AI-assisted technologies in the writing process

No AI tools/services were used during the preparation of this work.

## Acknowledgments

We sincerely express our gratitude to the homebound post-pancreaticoduodenectomy patients and primary caregivers who participated in this study and unreservedly shared their experiences and needs with us.

## Contributor Information

Qiuge Qiao.

Ci Dong.

